# Longitudinal changes in depressive symptoms following naturalistic ayahuasca use: A prospective cohort study in Las Vegas, Nevada

**DOI:** 10.64898/2026.08.24.26361280

**Authors:** Krista Whitley, Kai Castellarin, Kumar Dave, Taylor Parrott, Andre Craig Christie, Lisa Durette, Yusuf Khan, Kidus Yohannes, Rayan Muneer, Kristina Domanski

**Author notes:** Corresponding author: Krista Whitley, School of Global Public Health New York University, 708 Broadway, New York, NY, 10003 USA.

## Abstract

Ayahuasca use has expanded beyond its traditional Amazonian contexts, yet prospective longitudinal data examining depressive symptoms following naturalistic use in the United States remain limited. We conducted an interim analysis of an ongoing prospective observational cohort of adults participating in naturalistic ayahuasca use in Las Vegas, Nevada. Depressive symptoms were assessed using the Patient Health Questionnaire-9 (PHQ-9), with higher scores indicating greater depressive symptom severity, at baseline and approximately 48 hours, 30 days, 60 days, and 90 days following exposure. At interim analysis, PHQ-9 data were available for 87 participants at baseline, 61 at 48 hours, 41 at 30 days, 33 at 60 days, and 26 at 90 days. Mean PHQ-9 scores decreased from 8.06 (SD 5.96) at baseline to 4.39 at 48 hours, 3.76 at 30 days, 3.97 at 60 days, and 3.65 at 90 days. Among participants with matched baseline and follow-up assessments, mean changes were −3.58 points at 48 hours, −4.47 at 30 days, −4.48 at 60 days, and −4.76 at 90 days. In a mixed-effects model accounting for repeated observations, PHQ-9 scores remained significantly lower than baseline at 48 hours (β=−3.70; 95% CI −5.09 to −2.31), 30 days (β=−4.34; 95% CI −6.01 to −2.67), 60 days (β=−3.97; 95% CI −5.77 to −2.18), and 90 days (β=−4.25; 95% CI −6.18 to −2.33; all p<0.001). Among participants with baseline PHQ-9 scores ≥5 and matched follow-up data, 69.2% demonstrated a reduction of at least 5 points at 90 days. These interim findings provide preliminary evidence of a sustained longitudinal association between naturalistic ayahuasca exposure and lower depressive symptom scores through 90 days in a U.S.-based cohort. The observational design, self-selection, incomplete follow-up, and absence of a control group preclude causal inference. Continued longitudinal follow-up is needed to determine the durability of this association.

## Introduction

Depressive disorders are a major contributor to disability worldwide [1]. A substantial proportion of individuals continue to experience depressive symptoms despite available pharmacologic and psychotherapeutic treatments [2]. Growing interest in psychedelic compounds has therefore prompted investigation into their potential effects on depressive symptoms and other mental health outcomes. Ayahuasca, a psychoactive botanical preparation traditionally used by Indigenous communities of the Amazon basin, typically combines plants containing the serotonergic psychedelic N,N-dimethyltryptamine (DMT) with β-carboline-containing plants that inhibit monoamine oxidase and permit DMT to become orally active [3]. Although ayahuasca has longstanding ceremonial and spiritual uses in South America, its use has expanded internationally, including within religious and ceremonial settings in the United States [4].

Clinical research has identified a preliminary antidepressant signal following ayahuasca administration. In an early open-label study of six patients with recurrent depression, Osório et al. observed rapid reductions in depressive symptoms following a single dose of ayahuasca, with improvements measured using the Hamilton Depression Rating Scale and Montgomery–Åsberg Depression Rating Scale (MADRS) [5]. A subsequent double-blind randomized placebo-controlled trial involving 29 patients with treatment-resistant depression found significantly lower MADRS scores following ayahuasca compared with placebo, with between-group effects increasing through seven days after administration. At day 7, response occurred in 64% of participants receiving ayahuasca compared with 27% receiving placebo [6]. These findings provided controlled evidence that changes in depressive symptoms may occur rapidly following ayahuasca exposure while also raising an important question regarding whether observed improvements persist beyond the acute and early post-exposure periods.

Longer-term observational research has begun to address this question. Ruffell et al. prospectively followed 63 participants attending ceremonial ayahuasca retreats in the Peruvian Amazon and reported reductions in depressive symptoms and other measures of psychological distress that were sustained at six months [7]. In the Netherlands, van Oorsouw et al. followed 20 participants with clinically significant depressive symptoms before an ayahuasca-analogue ceremony and at subsequent assessments extending to one year. Mean Beck Depression Inventory scores decreased from 22.7 at baseline to 11.45 one day after the ceremony, 12.89 at one month, and 8.88 at one year; 17 participants remained in the study at the one-year assessment [8]. More recently, a longitudinal observational study of 280 adults in São Paulo state, Brazil, reported significant reductions in MADRS-measured depressive symptoms following ritualistic ayahuasca use, with improvements observed through 180 days of follow-up [9]. Together, these studies suggest that changes in depressive symptoms observed following ayahuasca exposure may extend beyond the immediate psychedelic experience and warrant continued longitudinal investigation [10].

Despite this expanding evidence base, important gaps remain. Much of the prospective literature examining ayahuasca and depression has been generated in Brazil, Peru, European settings, or international retreat environments. Studies have also used heterogeneous measures of depression, including the MADRS, Hamilton Depression Rating Scale, and Beck Depression Inventory, complicating comparisons across cohorts and limiting direct translation to commonly used U.S. clinical measures. The Patient Health Questionnaire-9 (PHQ-9) is a validated nine-item self-report measure widely used to assess depressive symptom severity in clinical and research settings [11]. However, prospective studies using repeated PHQ-9 measurements following naturalistic ayahuasca exposure remain comparatively limited.

Recent research involving U.S. military veterans illustrates both the growing interest in naturalistic psychedelic use and the remaining evidence gap. In a prospective study of 58 veterans attending naturalistic psychedelic retreats, including participants attending ayahuasca and psilocybin retreats, investigators observed significant improvements across multiple mental health outcomes, including PHQ-9 depressive symptom scores, approximately four weeks after the retreat [12]. Although these findings provide important evidence using the PHQ-9 in a U.S. population, psychedelic administration occurred in retreat settings and follow-up was limited to approximately one month. Longer prospective follow-up using repeated PHQ-9 measurements among individuals participating in naturalistic ayahuasca use in the United States remains limited.

Naturalistic studies provide an important complement to controlled clinical trials because ayahuasca use commonly occurs within ceremonial, religious, and community contexts rather than conventional medical settings. These environments may include social support, participant expectations, spiritual practices, preparation and integration activities, and other contextual factors capable of influencing mental health outcomes [13,14]. Consequently, observational studies cannot establish that changes in depressive symptoms are caused by ayahuasca itself.

Participant expectations, concurrent treatments, regression to the mean, selection effects, and measured or unmeasured characteristics of participants and the ceremonial environment may contribute to observed changes. Prospective longitudinal designs can nevertheless characterize the direction, magnitude, and persistence of symptom changes following real-world exposure and provide evidence to inform subsequent controlled research.

The present study reports an interim analysis of an ongoing prospective cohort study examining depressive symptoms among adults participating in naturalistic ayahuasca use in Las Vegas, Nevada. Depressive symptoms were assessed using the PHQ-9 before ayahuasca exposure and repeatedly during longitudinal follow-up. This interim analysis was undertaken to characterize the emerging trajectory of depressive symptoms among participants with available follow-up data, with particular attention to whether changes remained evident at 60 and 90 days following exposure. Given the ongoing nature of participant enrollment and follow-up, these findings are presented as an early longitudinal signal rather than evidence of treatment efficacy or a definitive estimate of long-term effect. Continued follow-up will allow assessment of the durability of these changes as the cohort matures.

The primary objective of this interim analysis was to characterize longitudinal changes in PHQ-9 depressive symptom scores following naturalistic ayahuasca exposure. Secondary objectives were to evaluate the magnitude and clinical relevance of observed changes across available follow-up periods and to characterize participant retention during the ongoing longitudinal study. We hypothesized that PHQ-9 scores would decrease following naturalistic ayahuasca exposure and that reductions would remain observable at 60- and 90-day follow-up among participants with available data.

## Materials and Methods

### Reporting guideline

**This manuscript was prepared in accordance with the Strengthening the Reporting of Observational Studies in Epidemiology (STROBE) statement for cohort studies.**

### Study design and setting

This study is an ongoing prospective observational cohort study examining longitudinal changes in mental health outcomes among adults participating in naturalistic ayahuasca use in Las Vegas, Nevada. The study was designed to characterize mental health outcomes associated with real-world ayahuasca participation rather than to evaluate ayahuasca as an investigator-administered therapeutic intervention. The research team did not assign participants to receive ayahuasca, determine the composition or dose of ayahuasca consumed, or administer ayahuasca as part of the research protocol.

Participants were recruited in collaboration with a community-based church in Las Vegas, Nevada, in which individuals independently elected to participate in ayahuasca ceremonies. Recruitment occurred through a study QR code posted within the church that allowed interested individuals to access the electronic study information and baseline survey directly. The church facilitated access to potential study participants, but participation in the research study was voluntary and separate from participation in ayahuasca ceremonies. Participant enrollment and longitudinal data collection began on September 1, 2025 and remain ongoing. The present interim analysis includes data accrued from September 1, 2025 through August 9, 2026.

### Ethical approval

The study protocol, participant surveys, consent materials, investigator documentation, and letter authorizing study conduct in collaboration with the study site were reviewed and approved by the University Medical Center of Southern Nevada Institutional Review Board (UMC IRB #UMC-2025-608) on August 20, 2025. The study received expedited review under 45 CFR 46.110(b)(1), Category 7, as minimal-risk research involving individual or group characteristics or behavior and survey methodology. The UMC IRB operates under Federalwide Assurance FWA#00002738 and is registered with the Office for Human Research Protections (IRB00002394). The study was conducted in accordance with applicable U.S. federal regulations governing the protection of human participants.

Participants reviewed an electronic consent statement before beginning the baseline survey and indicated informed consent by selecting “Yes” to proceed. A written signature was not collected. The UMC IRB determined that the study met the requirements for a Waiver of Documented Informed Consent because documented consent would be the only link to study participation, in accordance with 45 CFR 46.116(d). The IRB determined that the research involved no more than minimal risk to participants, that the waiver or alteration would not adversely affect participants’ rights or welfare, that the research could not practicably be carried out without the waiver or alteration, and that, whenever appropriate, participants would be provided with additional pertinent information after participation. Participation in the research was voluntary.

### Participants and recruitment

Individuals were eligible to participate if they were aged 18 years or older, independently planned to participate in naturalistic ayahuasca use, and had the capacity to provide informed consent. Individuals were excluded if they were younger than 18 years, lacked the capacity to provide informed consent, or were incarcerated or otherwise in custodial detention at the time of recruitment. Enrollment occurred before the participant’s ayahuasca experience, allowing collection of baseline mental health information before exposure and prospective follow-up thereafter.

Study participation did not determine eligibility for ayahuasca use or participation in a ceremony. Decisions regarding ceremonial participation occurred independently of the research team. Participants could discontinue research participation or decline subsequent surveys without affecting their relationship with the ceremonial organization.

### Longitudinal survey schedule

Participants were followed prospectively using repeated electronic surveys administered before and after naturalistic ayahuasca exposure. The study protocol includes assessments at baseline and approximately 48 hours, 30 days, 60 days, 90 days, 180 days, 365 days, and 720 days following exposure.

Because enrollment and longitudinal follow-up are ongoing, not all enrolled participants had reached every follow-up interval at the time of the interim analysis. Participants who had not yet reached a scheduled assessment were therefore distinguished from participants who had reached the assessment window but had not completed the corresponding survey. Specific reasons for nonresponse to scheduled follow-up surveys were not systematically collected. Later follow-up intervals will continue to accrue as the cohort matures.

### Baseline characteristics and exposure variables

Baseline demographic and exposure characteristics were collected through the electronic baseline survey before the participant’s ayahuasca experience. Variables included age, sex assigned at birth, gender identity, sexual orientation, self-reported race/ethnicity, military service, health insurance status, prior ayahuasca or other psychedelic use, and whether the upcoming ceremony represented the participant’s first ayahuasca experience.

### Depressive symptom assessment

The primary outcome for the present interim analysis was depressive symptom severity measured using the Patient Health Questionnaire-9 (PHQ-9). The PHQ-9 is a nine-item self-report instrument assessing the frequency of depressive symptoms during the preceding two weeks. Individual items are scored from 0 to 3, producing a total score ranging from 0 to 27, with higher scores indicating greater depressive symptom severity. Established severity categories were used descriptively: 0–4 minimal, 5–9 mild, 10–14 moderate, 15–19 moderately severe, and 20–27 severe depressive symptoms [11]. A PHQ-9 score of 10 or greater was used as an indicator of at least moderate depressive symptom burden.

### Narrative survey responses

In addition to structured mental health measures, longitudinal surveys included opportunities for participants to provide narrative information regarding their experiences, perceived changes in mental health and wellbeing, subsequent healthcare utilization, and other experiences occurring during follow-up.

Narrative responses were reviewed as contextual data to complement interpretation of quantitative PHQ-9 findings. Narrative responses were not treated as independent evidence of treatment efficacy and were not used to infer causality. Illustrative quotations were selected when they provided context for longitudinal quantitative findings or demonstrated meaningful variation in participant experiences.

To protect confidentiality, all narrative material presented in the manuscript was de-identified. Names, study numbers, email addresses, contact information, and other potentially identifying information were excluded from manuscript reporting.

### Interim analysis population

The present analysis represents an interim evaluation of an actively enrolling and ongoing longitudinal cohort. For registry follow-up accounting, a participant episode was defined as a ceremony-linked enrollment and follow-up sequence; an individual could contribute more than one participant episode if they enrolled around more than one ayahuasca ceremony during the study period. No a priori power-based sample-size calculation was performed for this interim analysis; the analytic sample was determined by the number of participants with available eligible data at the interim data cutoff. At the August 9, 2026 interim data cutoff, 87 participants had available baseline PHQ-9 data. Available PHQ-9 observations included 61 participants at approximately 48 hours, 41 at 30 days, 33 at 60 days, and 26 at 90 days.

Because participants entered the study at different times, absence of a later follow-up survey did not necessarily indicate loss to follow-up. Participants who had not yet reached a given assessment interval at the interim data cutoff were classified separately from participants for whom the assessment was due but no response had been received.

### Bias considerations

Several design and analytic procedures were used to reduce or assess potential sources of bias. Baseline PHQ-9 data were collected before naturalistic ayahuasca exposure, and research participation was kept separate from eligibility for or participation in ayahuasca ceremonies.

Participants who had not yet reached a scheduled follow-up interval were distinguished from participants whose assessment was due but not completed to avoid classifying ongoing cohort maturation as attrition. Within-participant matched analyses were used where possible, and baseline PHQ-9 scores were compared between later respondents and eligible nonrespondents to assess whether follow-up differed according to baseline depressive symptom severity. These procedures were intended to address or characterize potential bias but could not eliminate selection bias, expectancy effects, differential follow-up, or other measured and unmeasured confounding.

### Statistical analysis

Descriptive statistics were used to characterize PHQ-9 scores and participant follow-up across the interim study period. PHQ-9 scores were summarized at baseline and at approximately 48 hours, 30 days, 60 days, and 90 days following naturalistic ayahuasca exposure. For each assessment interval, the number of available observations, mean and standard deviation, median and interquartile range, and distribution of depressive symptom severity were calculated.

To evaluate within-participant changes in depressive symptoms, follow-up PHQ-9 observations were matched to each participant’s baseline PHQ-9 score when both measurements were available. Change scores were calculated by subtracting the baseline PHQ-9 score from the corresponding follow-up score, such that negative values represented reductions in depressive symptom severity. Mean within-participant changes and 95% confidence intervals were calculated separately for each follow-up interval.

The proportion of respondents with PHQ-9 scores of 10 or greater was calculated at each assessment, and within-participant transitions between severity categories were examined across available follow-up assessments.

Clinically meaningful change was evaluated using within-participant change from baseline. A reduction of at least 5 PHQ-9 points was used as a descriptive threshold based on prior PHQ-9 outcome research [15], and the proportion meeting this threshold was calculated at each follow-up interval among participants with matched baseline and follow-up measurements. Additional analyses examined the proportion demonstrating a reduction of at least 50% from baseline.

Because enrollment and longitudinal follow-up were ongoing at the time of this interim analysis, follow-up status was evaluated according to participant eligibility at each assessment interval.

Participants who had not yet reached a scheduled follow-up interval by the interim data cutoff were classified as not yet eligible and were not included in the denominator when calculating follow-up completion. Participants who had reached the corresponding follow-up window but had not completed the survey were classified separately as due with no response. This approach was used to distinguish ongoing cohort maturation from true nonresponse.

Follow-up completion was summarized at each assessment interval as the number of completed surveys divided by the number of participants who had reached that follow-up interval by the interim data cutoff. Patterns of missing follow-up data were examined descriptively, and baseline PHQ-9 scores were compared between participants with and without available later follow-up data to assess potential differences associated with attrition.

Given the repeated-measures structure of the study and unequal numbers of observations across follow-up intervals, longitudinal changes in PHQ-9 scores were additionally evaluated using a mixed-effects modeling approach incorporating available repeated observations. Time was modeled as a categorical fixed effect to allow changes in depressive symptoms to vary across assessment intervals, with participant included as a random intercept to account for within-participant correlation. The model was estimated using restricted maximum likelihood (REML) and included the 87 participants in the baseline PHQ-9 analytic cohort and their linked available follow-up PHQ-9 observations through the interim data cutoff (211 observations total). The model was not adjusted for participant-level demographic, clinical, or treatment covariates; estimates therefore represent unadjusted longitudinal associations. Estimated differences in PHQ-9 scores across follow-up intervals were reported with 95% confidence intervals. No additional sensitivity analyses were performed for this interim analysis. Data preparation was conducted in Microsoft Excel. Final statistical analyses were reproduced and verified using Python version 3.13.5, with NumPy version 2.3.5, SciPy version 1.17.0, and statsmodels version 0.14.6.

All analyses were interpreted within the context of an interim analysis of an ongoing observational cohort. Statistical findings were considered estimates of longitudinal associations and symptom trajectories rather than evidence of a causal treatment effect. No adjustment or statistical model can fully account for potential influences including self-selection, expectancy effects, ceremonial context, concurrent mental health treatment, social support, regression to the mean, differential follow-up, and other measured or unmeasured confounding factors.

## Results

The active registry contained 165 participant episodes. Because enrollment and longitudinal follow-up remained ongoing, participants who had not yet reached a scheduled assessment were distinguished from participants whose assessment was due but had not been completed. At approximately 48 hours, 61 of 164 participants who had reached the assessment window had completed the survey (37.2%), with one participant not yet due. At 30 days, 41 of 165 eligible participants had completed the assessment (24.8%). At 60 days, 33 of 154 participants who had reached the assessment window had completed the survey (21.4%), while 11 participants were not yet due. At 90 days, 26 of 144 participants who had reached the assessment window had completed the survey (18.1%), while 21 participants had not yet reached the 90-day assessment. Follow-up status across the active registry is summarized in Fig 1.

**Fig 1.**
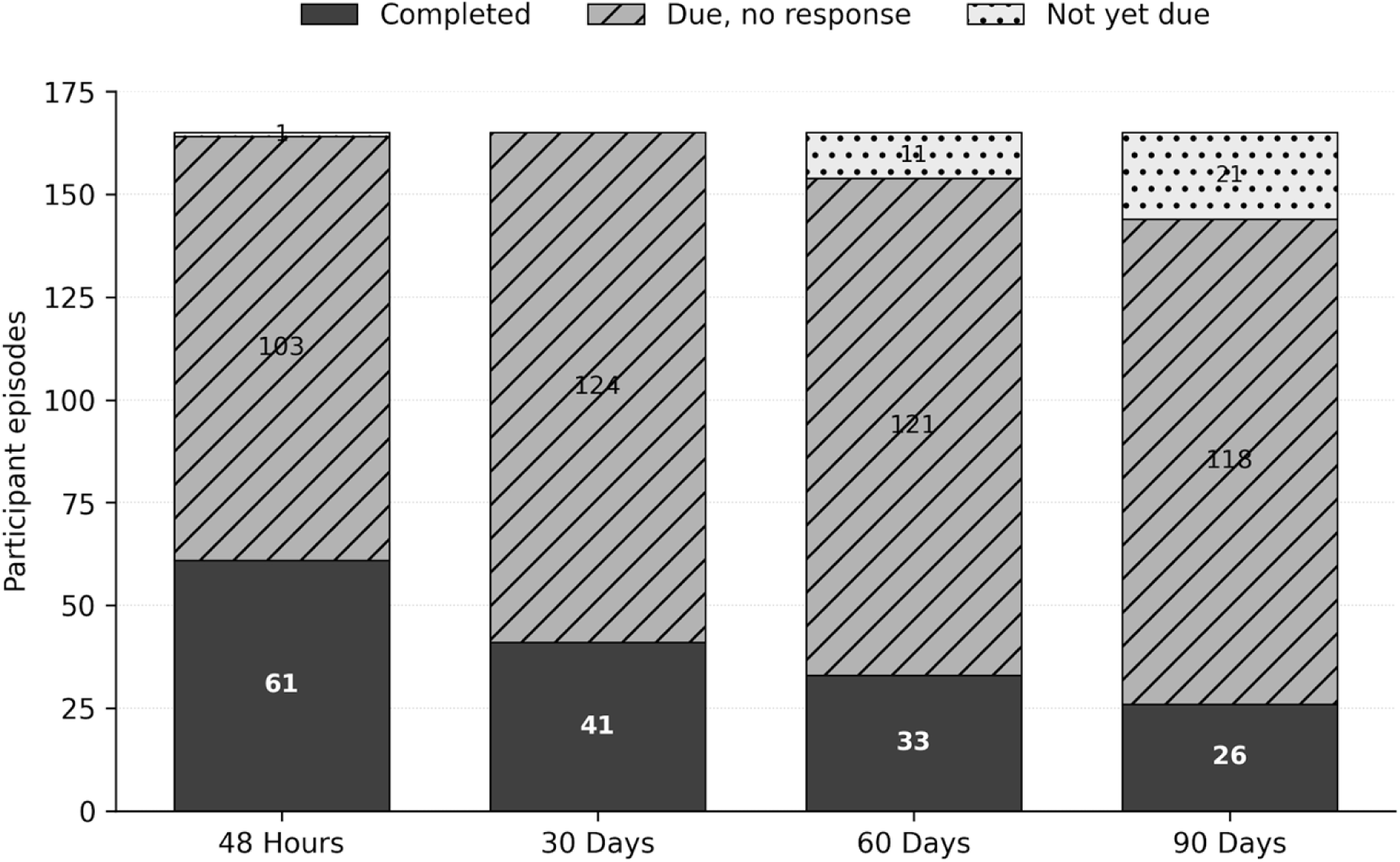
Study follow-up status across assessment intervals in the active registry. Bars show completed surveys, participant episodes that had reached the assessment window but had not responded, and participant episodes that were not yet due at the interim data cutoff. Registry follow-up status is distinct from the subset of participants contributing PHQ-9 data to the depressive symptom analyses.

### Longitudinal PHQ-9 scores

PHQ-9 scores were lower than baseline at each available follow-up assessment. At baseline, the mean PHQ-9 score was 8.06 (SD 5.96; median 8, IQR 3–11; n=87). The mean score decreased to 4.39 at approximately 48 hours (SD 5.44; median 3, IQR 1–5; n=61) and 3.76 at 30 days (SD 4.39; median 3, IQR 1–5; n=41). Mean scores remained lower at 60 days (mean 3.97, SD 3.70; median 3, IQR 1–6; n=33) and 90 days (mean 3.65, SD 4.07; median 2, IQR 1–6; n=26).

The wave-specific means therefore demonstrated a substantial early decline in depressive symptom scores followed by relatively stable mean PHQ-9 scores from 30 through 90 days. Because the participants contributing data differed across assessment intervals, these cross-sectional estimates were supplemented with within-participant and longitudinal mixed-effects analyses. Wave-specific means and confidence intervals are shown in Fig 2, and descriptive outcomes are summarized in Table 1.

**Fig 2.**
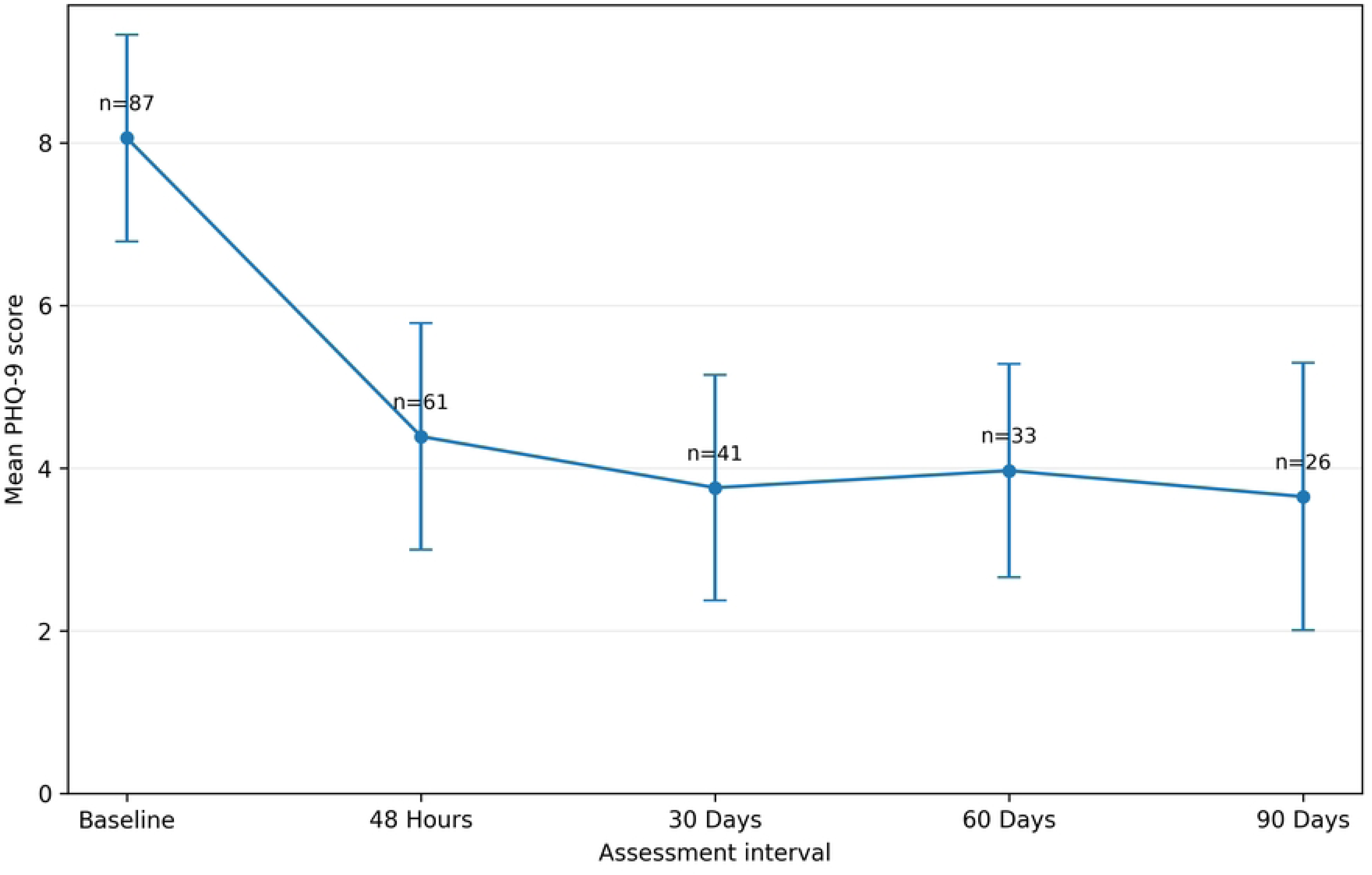
Mean PHQ-9 scores from baseline through 90 days following naturalistic ayahuasca exposure. Error bars represent 95% confidence intervals for the wave-specific mean; sample size is shown at each assessment interval.

**Table 1.** PHQ-9 outcomes by assessment interval.

| Assessment | n | Mean (SD) | Median (IQR) | PHQ-9 ≥10, n (%) |
| --- | --- | --- | --- | --- |
| Baseline | 87 | 8.06 (5.96) | 8 (3–11) | 33/87 (37.9%) |
| 48 Hours | 61 | 4.39 (5.44) | 3 (1–5) | 9/61 (14.8%) |
| 30 Days | 41 | 3.76 (4.39) | 3 (1–5) | 4/41 (9.8%) |
| 60 Days | 33 | 3.97 (3.70) | 3 (1–6) | 5/33 (15.2%) |
| 90 Days | 26 | 3.65 (4.07) | 2 (1–6) | 2/26 (7.7%) |

### Baseline characteristics

**Baseline demographic and exposure characteristics of the 87 participants contributing baseline PHQ-9 data are summarized in Table 2. Participants were represented across all age categories shown, with the largest group aged 41–45 years (19/87, 21.8%); 46 (52.9%) were female by sex assigned at birth, 60 (69.0%) self-identified as White, 12 (13.8%) reported a history of military service, and 73 (83.9%) reported prior ayahuasca or other psychedelic use.**

**Table 2.** Baseline characteristics of the PHQ-9 analytic cohort (n=87).

| **Characteristic** | **Category** | **n (%)** |
| --- | --- | --- |
| **Age group, years** | 18-25 | 1 (1.1%) |
|  | 26-30 | 2 (2.3%) |
|  | 31-35 | 11 (12.6%) |
|  | 36-40 | 13 (14.9%) |
|  | 41-45 | 19 (21.8%) |
|  | 46-50 | 13 (14.9%) |
|  | 51-55 | 14 (16.1%) |
|  | 56-60 | 8 (9.2%) |
|  | 61-65 | 4 (4.6%) |
|  | 66-70 | 1 (1.1%) |
|  | 71-75 | 1 (1.1%) |
| **Sex assigned at birth** | Female | 46 (52.9%) |
|  | Male | 41 (47.1%) |
| **Gender identity** | Female | 46 (52.9%) |
|  | Male | 40 (46.0%) |
|  | Prefer not to say | 1 (1.1%) |
| **Sexual orientation** | Heterosexual / Straight | 68 (78.2%) |
|  | Asexual | 7 (8.0%) |
|  | Bisexual | 4 (4.6%) |
|  | Gay | 2 (2.3%) |
|  | Prefer not to say | 2 (2.3%) |
|  | Queer | 2 (2.3%) |
|  | Lesbian | 1 (1.1%) |
|  | Questioning | 1 (1.1%) |
| **Race/ethnicity\*** | White | 60 (69.0%) |
|  | Hispanic/Latino/a/x or Tejano | 10 (11.5%) |
|  | Asian | 6 (6.9%) |
|  | Black or African American | 6 (6.9%) |
|  | Native Hawaiian or Other Pacific Islander | 2 (2.3%) |
|  | Multiracial/multiethnic | 2 (2.3%) |
|  | Other self-description | 1 (1.1%) |
| **Military service** | Yes | 12 (13.8%) |
|  | No | 75 (86.2%) |
| **Health insurance** | Yes | 67 (77.0%) |
|  | No | 20 (23.0%) |
| **Prior ayahuasca/psychedelic use** | Yes | 73 (83.9%) |
|  | No | 14 (16.1%) |
| **First time using ayahuasca** | Yes | 47 (54.0%) |
|  | No | 40 (46.0%) |

*\* Race/ethnicity categories were harmonized from self-reported responses. Tejano was grouped with Hispanic/Latino/a/x; White/Asian and Black American/Mexican American responses were grouped as multiracial/multiethnic. Percentages may not sum to 100 due to rounding.*

### Within-participant changes from baseline

Among participants with both baseline and follow-up PHQ-9 measurements, depressive symptom scores were lower than baseline at every follow-up interval.

At approximately 48 hours, the mean within-participant change was −3.58 points (SD 5.22; 95% CI −5.10 to −2.07; n=48; p<0.001). At 30 days, the mean change was −4.47 points (SD 5.49; 95% CI −6.52 to −2.42; n=30; p<0.001). At 60 days, the mean change was −4.48 points (SD 6.04; 95% CI −6.97 to −1.99; n=25; p=0.001). At 90 days, the mean within-participant change was −4.76 points (SD 5.17; 95% CI −7.11 to −2.41; n=21; p<0.001).

The magnitude of the mean within-participant reduction was therefore similar at the 30-, 60-, and 90-day assessments, with no apparent attenuation toward baseline during this interval among participants with available matched data.

### Mixed-effects longitudinal analysis

A mixed-effects model was used to evaluate repeated PHQ-9 observations within the baseline analytic cohort while accounting for within-participant correlation and unequal numbers of observations across follow-up intervals. The model included 211 observations: 87 baseline observations and 124 linked follow-up observations from those same participants (48 at approximately 48 hours, 30 at 30 days, 25 at 60 days, and 21 at 90 days). Time was modeled as a categorical fixed effect and participant was included as a random intercept.

Relative to baseline, estimated PHQ-9 scores were 3.70 points lower at approximately 48 hours (β=−3.70; 95% CI −5.09 to −2.31; p<0.001), 4.34 points lower at 30 days (β=−4.34; 95% CI −6.01 to −2.67; p<0.001), 3.97 points lower at 60 days (β=−3.97; 95% CI −5.77 to −2.18; p<0.001), and 4.25 points lower at 90 days (β=−4.25; 95% CI −6.18 to −2.33; p<0.001).

The mixed-effects estimates were consistent with the descriptive and matched analyses, demonstrating lower depressive symptom scores at each available follow-up interval and persistence of the observed difference through 90 days.

### Clinically meaningful change

A reduction of at least 5 PHQ-9 points was examined at the individual level. Among all participants with matched baseline and follow-up data, this threshold was met by 18 of 48 participants (37.5%) at approximately 48 hours, 12 of 30 (40.0%) at 30 days, 11 of 25 (44.0%) at 60 days, and 9 of 21 (42.9%) at 90 days.

Because participants with baseline PHQ-9 scores below 5 could not achieve a 5-point decrease, this outcome was also examined among participants with baseline scores of at least 5. Within this group, 18 of 32 participants (56.3%) demonstrated a reduction of at least 5 points at approximately 48 hours, 12 of 20 (60.0%) at 30 days, 11 of 16 (68.8%) at 60 days, and 9 of 13 (69.2%) at 90 days.

Among participants with baseline PHQ-9 scores greater than zero and matched follow-up measurements, a reduction of at least 50% from baseline was observed in 30 of 47 participants (63.8%) at approximately 48 hours, 19 of 30 (63.3%) at 30 days, 15 of 25 (60.0%) at 60 days, and 15 of 21 (71.4%) at 90 days. Matched and longitudinal outcomes are summarized in Table 3.

**Table 3.** Matched and longitudinal PHQ-9 outcomes following naturalistic ayahuasca exposure.

| Outcome | 48 Hours | 30 Days | 60 Days | 90 Days |
| --- | --- | --- | --- | --- |
| Matched n | 48 | 30 | 25 | 21 |
| Matched mean change (95% CI) | -3.58 (-5.10, -2.07) | -4.47 (-6.52, -2.42) | -4.48 (-6.97, -1.99) | -4.76 (-7.11, -2.41) |
| Mixed-effects $\beta$ (95% CI) | -3.70 (-5.09, -2.31) | -4.34 (-6.01, -2.67) | -3.97 (-5.77, -2.18) | -4.25 (-6.18, -2.33) |
| $\geq 5$-point reduction† | 56.3% (18/32) | 60.0% (12/20) | 68.8% (11/16) | 69.2% (9/13) |
| $\geq 50\%$ | 63.8% (30/47) | 63.3% (19/30) | 60.0% (15/25) | 71.4% (15/21) |
| reduction* |  |  |  |  |

† Among participants with baseline PHQ-9 $\geq 5$ and matched follow-up data. \* Among participants with baseline PHQ-9 $> 0$ and matched follow-up data.

### Depressive symptom burden

At baseline, 33 of 87 participants (37.9%) had a PHQ-9 score of 10 or greater, corresponding to at least moderate depressive symptom burden. The proportion of respondents meeting this threshold was lower at every subsequent assessment: 9 of 61 (14.8%) at approximately 48 hours, 4 of 41 (9.8%) at 30 days, 5 of 33 (15.2%) at 60 days, and 2 of 26 (7.7%) at 90 days.

The distribution of PHQ-9 severity categories also shifted toward lower symptom levels over follow-up. At baseline, 31 participants had minimal symptoms (PHQ-9 0–4), 23 had mild symptoms (5–9), 22 had moderate symptoms (10–14), 5 had moderately severe symptoms (15–19), and 6 had severe symptoms (20–27). At 90 days, among the 26 respondents with available PHQ-9 data, 17 had minimal symptoms (0–4), 7 had mild symptoms (5–9), 1 had moderate symptoms (10–14), and 1 had moderately severe symptoms (15–19); no respondent had a PHQ-9 score in the severe range (20–27).

### Within-participant severity transitions

Changes in PHQ-9 severity category were examined among participants with matched baseline and follow-up assessments. At approximately 48 hours, 21 of 48 participants (43.8%) moved to a lower depressive symptom severity category, 23 (47.9%) remained in the same category, and 4 (8.3%) moved to a higher category.

At 30 days, 17 of 30 participants (56.7%) moved to a lower severity category, 12 (40.0%) remained unchanged, and 1 (3.3%) moved to a higher category. At 60 days, 13 of 25 participants (52.0%) were in a lower severity category than at baseline, 10 (40.0%) remained in the same category, and 2 (8.0%) moved to a higher category. At 90 days, 10 of 21 participants (47.6%) were in a lower severity category and 11 (52.4%) remained in the same category; none of the participants with matched 90-day data were in a higher PHQ-9 severity category than at baseline.

### Individual variability

Individual trajectories varied despite lower cohort-level mean scores. Some participants demonstrated substantial reductions, whereas others showed little change or higher PHQ-9 scores at later assessments, and moderate or greater symptom burden remained present among some respondents at 60 and 90 days. These findings demonstrate heterogeneity but do not establish that participants with higher baseline severity were more likely to remain symptomatic.

### Assessment of differential follow-up

Baseline PHQ-9 scores were compared between participants who completed later follow-up assessments and participants who had reached the corresponding assessment window but did not respond.

At the 48-hour assessment, the mean baseline PHQ-9 score was 7.92 among later respondents and 8.18 among eligible nonrespondents (p=0.84). At 30 days, baseline means were 8.03 among respondents and 8.07 among nonrespondents (p=0.98). At 60 days, baseline means were 8.36 and 7.90, respectively (p=0.76), and at 90 days were 8.29 and 7.94, respectively (p=0.83).

No statistically significant differences in baseline PHQ-9 scores were identified between respondents and eligible nonrespondents at any follow-up interval. These findings provide some reassurance that later responders were not selected solely on the basis of baseline depressive symptom severity; however, differential follow-up according to other measured or unmeasured participant characteristics remains possible.

### Participant-reported experiences

Narrative survey responses provided contextual information regarding participants’ experiences during longitudinal follow-up. Several participants described perceived improvements in mood, emotional processing, or general wellbeing that occurred alongside lower PHQ-9 scores.

One participant reported at the 30-day assessment, “It’s been more than 30 days I feel so much better. I feel like I am more open to life,” and had a PHQ-9 score of 3 at that assessment.

At the 60-day assessment, another participant described “processing and moving through my thoughts and feelings with ease” and had a PHQ-9 score of 2.

Narrative responses were heterogeneous and were considered illustrative contextual data rather than independent evidence of therapeutic efficacy. Participant narratives were not used to attribute changes in depressive symptoms specifically to ayahuasca.

## Discussion

### Principal findings

In this interim prospective cohort, PHQ-9 scores were lower than baseline at every available follow-up through 90 days. The pattern remained evident in matched analyses, with mean within-participant reductions of 4.47 points at 30 days, 4.48 at 60 days, and 4.76 at 90 days. These estimates suggest persistence of the observed association through three months among participants with available data, while the observational design precludes causal inference.

The proportion of respondents with PHQ-9 scores ≥10 was likewise lower at each follow-up than at baseline, although these wave-specific proportions should be interpreted cautiously because respondent composition changed over time.

### Comparison with previous research

The trajectory observed here is broadly consistent with prior clinical and naturalistic research. Osório et al. reported rapid reductions in depression scores after a single ayahuasca administration among six patients with recurrent depression [5], and Palhano-Fontes et al. found greater MADRS reductions with ayahuasca than placebo among 29 patients with treatment-resistant depression [6]. Those studies primarily addressed acute and early post-administration outcomes.

Longer naturalistic studies have reported sustained changes: Ruffell et al. observed improvements through six months in Peruvian retreat participants [7], van Oorsouw et al. reported lower Beck Depression Inventory scores among participants retained to one year in the Netherlands [8], and Camargos et al. reported lower MADRS-measured symptoms through 180 days in Brazil [9].

The present study adds repeated PHQ-9 measurements in a civilian U.S. cohort specifically participating in naturalistic ayahuasca use. Calnan et al. reported PHQ-9 improvements approximately four weeks after naturalistic psychedelic retreats among 58 U.S. military veterans [12]; the current cohort extends PHQ-9 follow-up to 90 days in a different U.S. population.

### Persistence of depressive symptom changes

The relative stability of matched PHQ-9 reductions from 30 through 90 days is notable, although later estimates are based on progressively smaller samples and cannot establish a treatment effect. Reductions approached five points, a threshold used in prior PHQ-9 outcome research [15], but mean change can obscure individual variability. Planned 180-, 365-, and 720-day assessments will help determine whether the observed trajectory persists or attenuates and whether it differs by baseline symptom burden or other participant characteristics.

### Naturalistic context and interpretation

The naturalistic design is both a strength and a limitation. Participants independently chose ceremonial ayahuasca use, and the research team neither assigned nor standardized the exposure. Preparation, expectations, social support, spiritual meaning, facilitator and peer interactions, and post-ceremony integration may all influence mental health outcomes [13,14]; these contextual components cannot be separated from the pharmacologic exposure in this study.

Narrative responses provided context for how some participants perceived changes in mood or emotional processing, but they were not analyzed as an independent outcome and do not establish that ayahuasca produced those experiences. PHQ-9 trajectories were also heterogeneous, with clinically relevant symptoms persisting among some respondents; future analyses should examine characteristics associated with differing trajectories.

### Relevance of the United States setting

The U.S. setting addresses a geographic gap in prospective ayahuasca research, much of which has been conducted in South America, Europe, or international retreat environments. Studying a Las Vegas cohort prospectively while using the widely implemented PHQ-9 [11] provides data that may be more directly comparable with other U.S. mental health research, while still requiring caution about generalizability.

### Strengths and limitations

Strengths include prospective baseline assessment before exposure, repeated longitudinal follow-up, use of the validated PHQ-9, and a U.S.-based naturalistic cohort. Narrative responses provided limited contextual information alongside the quantitative measures.

Several limitations are also important. Most importantly, this was an observational study without a control group, and causal conclusions cannot be drawn. Participants self-selected both into naturalistic ayahuasca use and into research participation, introducing the potential for selection bias. In addition, 83.9% of the baseline PHQ-9 analytic cohort reported prior ayahuasca or other psychedelic use, which may limit generalizability to psychedelic-naïve populations and may also influence participant expectations surrounding the experience. Expectancy effects, ceremonial and religious context, social support, concurrent mental health treatment, changes in medication, life events, regression to the mean, and other measured or unmeasured factors may have contributed to changes in PHQ-9 scores.

The naturalistic design also meant that the research team did not control or standardize the composition or dose of ayahuasca consumed. Consequently, the study cannot evaluate dose-response relationships or attribute observed outcomes to a standardized pharmacologic exposure.

Attrition and incomplete follow-up represent additional limitations. Sample sizes decreased across later assessments, and participants who continued responding may differ systematically from those who did not. Because the study remains ongoing, however, not all missing later assessments represent attrition; some participants had not yet reached the corresponding follow-up interval at the interim data cutoff. Later-wave findings are therefore based on smaller samples and should be considered preliminary. Baseline PHQ-9 scores did not differ significantly between later respondents and eligible nonrespondents at any assessed interval; however, continued follow-up and comparisons using additional participant characteristics will remain important for evaluating potential attrition bias.

Self-reported PHQ-9 scores are also subject to reporting bias and do not constitute a clinical diagnosis of major depressive disorder. The cohort included participants across a range of baseline depressive symptom severity rather than exclusively individuals with diagnosed depressive disorders. Accordingly, these findings should be interpreted as changes in self-reported depressive symptoms within a naturalistic cohort rather than evidence regarding treatment of major depressive disorder.

### Implications and future research

The persistence of lower PHQ-9 scores through 90 days supports follow-up beyond acute and short-term outcomes. Continued assessment through 180, 365, and 720 days should clarify durability, patterns of missingness, and participant characteristics associated with differing trajectories. Controlled research will still be necessary to distinguish pharmacologic effects from expectancy, ceremonial context, social support, and other components of the naturalistic experience; prospective real-world studies can complement that work by characterizing outcomes in settings where ayahuasca is actually used.

### Conclusion

In this interim U.S. cohort, depressive symptom scores were lower following naturalistic ayahuasca exposure and remained below baseline through 90 days among participants with available follow-up data. Because the study is observational, self-selected, and lacks a control group, these findings describe an association rather than a causal effect. Continued follow-up through 180, 365, and 720 days will help characterize durability, individual variability, and factors associated with differing symptom trajectories.

## Author contributions

**Krista Whitley: Conceptualization, Investigation, Methodology, Project administration, Supervision, Writing – original draft, Writing – review & editing.**

**Kai Castellarin: Data curation, Investigation, Methodology, Project administration. Kumar Dave: Conceptualization, Writing – original draft.**

**Taylor Parrott: Formal analysis, Investigation.**

**Andre Craig Christie: Project administration, Writing – review & editing. Lisa Durette: Validation, Writing – review & editing.**

**Yusuf Khan: Methodology.**

**Kidus Yohannes: Project administration, Visualization, Writing – original draft. Rayan Muneer: Investigation, Project administration, Writing – review & editing. Kristina Domanski: Validation, Writing – review & editing.**

## Acknowledgments

The authors thank the participants who contributed their time and experiences to this ongoing longitudinal study. The authors also acknowledge the collaborating community-based study site for facilitating access to individuals interested in participating in the research and supporting study recruitment. Participation in the research remained voluntary and separate from participation in ayahuasca ceremonies, and the research team retained responsibility for data collection, analysis, interpretation, and manuscript preparation.

## Data Availability

Data supporting the findings reported in this manuscript are provided within the manuscript and its Supporting Information files. The de-identified participant-level longitudinal PHQ-9 dataset used for the primary quantitative analyses, together with a variable codebook, is available as S1 Dataset. Aggregate demographic and study follow-up data are reported in the manuscript tables and text. Reproducibility code for the principal quantitative analyses is available as S2 Code.

## Supporting Information

S1 Dataset. De-identified longitudinal PHQ-9 dataset used for the analyses reported in this study, including a variable codebook.

S2 Code. Python code used to reproduce the principal descriptive, matched within-participant, severity-transition, clinically meaningful change, and mixed-effects analyses reported in the manuscript.

